# Sleep disorders predict the one-year onset, persistence, but not remission of psychotic experiences in pre-adolescence: a longitudinal analysis of the ABCD cohort data

**DOI:** 10.1101/2021.01.15.21249864

**Authors:** Sarah Reeve, Vaughan Bell

## Abstract

The relationship between sleep disorder and psychotic experiences in preadolescence has not been extensively studied despite the potential for intervention. The current study addresses this relationship using the Adolescent Brain Cognitive Development (ABCD) cohort, which provides baseline data from 11,830 10 to 11-year-olds; for 4910 of these 1-year follow-up data is also available. A set of pre-registered multi-level regression models were applied to test whether a) sleep disorder is associated with psychotic experiences at baseline; b) baseline sleep disorder predicts psychotic experiences at follow-up; c) the persistence of sleep disorder predicts persistence of psychotic experiences at follow-up; d) the remission of sleep disorder predicts the remission of psychotic experiences at follow-up. After controlling for potential confounders, sleep disorder was associated with psychotic experiences cross-sectionally (OR=1.40, 95% CI 1.20-1.63), at one-year follow-up (OR=1.32, 95% CI 1.11-1.57), and the persistence of sleep disorder predicted the persistence of psychotic experiences (OR=1.72, 95% CI 1.44-2.04). However, remission of sleep problems did not predict remission of psychotic experiences (OR=1.041, 95% CI 0.80-1.35). The results indicate that sleep disorders in preadolescence are common and associated with psychotic experiences, although the lack of co-remission raises questions about the mechanism of association. However, given these findings, and existing evidence in later adolescence and adults, further investigation of sleep as a preventative mental health intervention in this age group is warranted.

## Introduction

Sleep disorders are hypothesised to be a significant causal factor in the development and maintenance of psychosis [1, 2]. Experimental research in adults supports that sleep disruption increases psychotic experiences [3], and that treating sleep disorders reduces psychotic experiences in adults [4]. While the relationship between sleep and psychotic symptoms is thought to be bidirectional, longitudinal studies in adults have indicated that sleep disorders predict later psychotic symptoms to a greater extent than vice versa [5, 6]. These and other findings have raised the possibility of targeting sleep disorders as a preventative, early intervention in mental health [4, 7, 8].

It is known that psychotic-like experiences are roughly 10x more common in children than they are in adults (reported by 66% of 7-8 year old children compared to 5.8-7.2% of adults; [9–11]. Not all childhood psychotic experiences are pathological [12] nevertheless persistence, distress, and amount of different psychotic experiences reported in childhood is associated with higher risk of developing adult psychotic disorder [13, 14] and a predictor of poor physical and mental health more generally later in life [15–17]. The age at which psychotic experiences are reported is also positively related to risk of later psychotic disorder. Reporting psychotic experiences at age 10-11 is associated with a 5x increased likelihood of developing a later psychotic disorder [14], whereas reporting these experiences by ages 14-17 is associated with a 10x increased likelihood [13]. This set of findings have been encompassed under a ‘proneness to persistence’ model of psychosis, with these non-clinical psychotic experiences existing on a continuum with psychotic disorders [10].

The relationship between sleep disorders and psychotic experiences in children is less well-researched than in adults [1]. However, many studies have demonstrated that children or adolescents with sleep disturbances are more likely to report psychotic experiences [18–20]. Several large population cohort studies have reported that sleep problems during childhood predict psychotic experiences contemporaneously and across adolescence [19, 21–23]. Furthermore, as in adults, sleep problems in children are linked to a wide range of other negative mental and physical health outcomes [24–26].

Despite this evidence indicating that sleep disorders have an important role in the onset of psychotic experiences during childhood and adolescence, it is less clear whether sleep disorders contribute to persistence of psychotic experiences in this age group. This is particularly important given that persistence is a particularly strong predictor of poor outcome. It is also notable that a high proportion of both sleep problems and psychotic experiences remit during adolescence [27] – yet as with persistence, whether sleep problems have an influence on remission of psychotic experiences is unknown. The large cohort studies listed above have rarely measured both sleep and psychotic experiences at multiple time points (with sleep being assessed only at younger ages and psychotic experiences assessed only assessed in teenage years) making it difficult to explore their concurrent relationship.

### The current study

We used the Adolescent Brain and Cognitive Development study cohort data (release 2.0.1) to investigate the relationship between sleep and psychotic experiences in a cohort of 9-11 year-olds to address a set of pre-registered hypotheses:

1. Do sleep disorders and psychotic experiences co-occur in 9-10 year old children?
2. Do sleep disorders predict later onset of psychotic experiences?
3. Do sleep disorders predict persistence of psychotic experiences?
4. Does remission of sleep disorders predict remission of psychotic experiences?

These were examined use a pre-registered set of linear and logistic regression models applied to the baseline and one-year follow up data using a pre-registered analysis plan (https://osf.io/8ks72/)

## Method

### Recruitment

The Adolescent Brain and Cognitive Development (ABCD; https://abcdstudy.org/) 2.0.1 release study data was used in the current analysis [28, 29]. The ABCD study is an ongoing longitudinal cohort study aimed at recruiting a representative sample of US children. At the initial stage, all families of children aged 9-10 in geographic catchment area of study sites across the USA were contacted via schools with information about the study. Volunteer families were then screened for inclusion. Participants were purposively recruited to match national US sociodemographic factors, for example utilising targets for each of five major race/ethnicity classifications (White, African-American, Hispanic, Asian All Other). The 2.0.1 release includes baseline data on 11,873 individuals, and one year follow-up from 4,951 of those participants, with follow up data from the remainder of the cohort to be released at a future date. Full details of the ABCD study design are available in a special issue of Developmental Cognitive Neuroscience [28].

### Measures

#### Psychotic experiences

Participants’ psychotic experiences were assessed using the Prodromal Questionnaire – Brief Child version (PQ-BC). This measure was validated within the current ABCD dataset showing high reliability (Cronbach’s alpha = 0.863 for total score and 0.873 for distress subscale; [30]. The PQ-BC is a self-report questionnaire assessing presence and distress associated with psychotic like symptoms in children. For each psychotic experience the child is asked if they experience it (yes/no), and if they do, how much it distresses/bothers them on a pictographic 1 to 5 scale showing a human cartoon figure in various levels of distress. This questionnaire yields two outcome variables – the sum of symptoms endorsed (range 0-21) and the sum of distress reported for those symptoms (range 0-126).

For the current study, both continuous and dichotomous outcomes of the sum of symptoms were used. For the dichotomous outcome the symptom score was transformed to a categorical 0-1 variable where 1 indicates presence of at least one psychotic symptom and 0 indicates no psychotic symptoms present. When applied to baseline and follow up this was then used to derive variables indicating ‘new onset’ of psychotic symptoms (i.e. 0 at baseline, 1 at follow up) ‘persistence’ of psychotic symptoms (i.e. 1 at both time points), and ‘remission’ (1 at baseline, 0 at follow up).

As an addition to our analysis plan (see Alterations to the pre-registered statistical plan for further details and rationale) we derived a count of *distressing* psychotic experiences which was dichotomised to represent the presence/absence of at least one distressing psychotic symptom (≥2 score on distress rating).

### Sleep disorders

Presence of sleep disorders was assessed using the Sleep Disorder Scale for Children (SDSC), which is a parent-reported questionnaire assessing the presence of a range of sleep disorder symptoms in children [31]. It is composed of 26 Likert items assessing the frequency of various disturbances over the past 6 months on a 1 to 5 Likert scale (1 = never, 5 = always/daily experiencing a particular issue). The total score provides a measure of sleep disturbance, for which a cut-off point at 39 has sensitivity of 0.89 and specificity of 0.74, correctly identifying 73.4% of a control group and 89.1% of sleep disordered participants.

This cut-off was used to categorise participants according to absence or presence of disturbed sleep. The categorical score at baseline and follow up was then used to derive variables indicating ‘persistence’ of sleep disturbance (i.e. present at both time points), ‘remission’ (present at baseline, absent at follow up), and ‘onset’ (absent at baseline, present at follow up)

### Potential confounders: Socio-demographic, IQ, and medication variables

Further variables were used from the ABCD dataset to index potential confounders, defined as factors that can independently influence each of the variables of interest (sleep and psychotic experiences).

- Male gender and non-white ethnicity are associated with a higher likelihood of reporting both sleep problems [32] and psychotic experiences [33]. Gender (Male/Female), ethnicity (White/Black/Hispanic/Asian/Other) were reported within the basic demographic questionnaires of the study. Ethnicity was re-coded in to white/non-white for the purposes of all analyses.
- Lower socioeconomic status is also associated with increased likelihood of psychiatric disorder [34] and shorter sleep duration [35]. Socioeconomic status was indexed by using the sum score (range = 0-7) of seven yes/no items in the parent demographic survey questions relating to experiences of family hardship (e.g. “*in the past 12 months has there been a time when you and your immediate family needed food but couldn*’*t afford to buy it or couldn*’*t afford to go out to get it?*”), with higher scores on this sum scale indicating lower socioeconomic status. Neighbourhood deprivation was also assessed using the area deprivation index of the home address, which provides a national percentile value (range = 1-100) with higher values indicating higher levels of deprivation.
- Family conflict is also associated with sleep problems [36] and psychotic experiences [37]. Family conflict was indexed by the 9-item family conflict subscale of the family experiences. Each item is reported by parents as true or false (e.g. “*We fight a lot in our family*”), with higher values indicating higher levels of conflict (range = 0-9).
- Lower IQ scores and prescription of stimulant medications have been reported to have associations with psychotic experiences [38] and, especially for stimulant medications, with sleep problems [39]. Child IQ was assessed using the WISC-V matrix reasoning subscale score (range = 1-19), with higher values indicating higher IQ. Medication fields were searched for any stimulant medications (e.g. “*Methylphenidate*”) and their trade names (e.g. “*Ritalin*”) with absence or presence coded as a dichotomous 0/1 variable.

Notably, depression and anxiety were not included as potential confounders as these are consistently found to act as mediators in the causal pathway between sleep and psychotic experiences (e.g. [3]). Therefore, if included in statistical models as confounders this would likely result in an underestimate of the relationship between sleep and psychotic experiences which was the primary focus of the current investigation.

### Analysis

The pre-registration document and the analysis code used in this study are available online at the following link: https://osf.io/8ks72/. R version 3.6.2 was used for all analyses. A list of packages and version numbers used can be found in Supplementary Material 1. The pre-registration was completed before the ABCD 2.0 data release, i.e. before the one-year follow-up data was made available.

For each research question a set of planned regression analyses were pre-specified. In each case, the regression model was first estimated with only the key explanatory (sleep) and dependent (psychosis) variables. If a significant association was found, potential confounders were added to test robustness of the hypothesised association.

The following four research questions were tested:

#### 1. Do sleep disorders and psychotic experiences co-occur?

This was investigated cross-sectionally within baseline using a) a linear regression to test continuous association between sleep symptoms and psychotic experiences b) a logistic regression to test if sleep symptoms (continuous) predicted presence of psychotic experiences (dichotomous) and c) a logistic regression to test if presence of sleep disorder (dichotomous) predicted presence of psychotic experiences (dichotomous).

#### 2. Do sleep disorders predict later psychotic experiences?

This was investigated by logistic regression models testing if the presence of sleep disorder a) at baseline and b) at both baseline and follow-up predicted psychotic experiences at one year follow up.

#### 3. Do sleep disorders predict persistence of psychotic experiences?

This was investigated by logistic regression models testing if the presence or persistence of sleep disorder predicted persistence (i.e. presence at both baseline and follow-up) of psychotic experiences.

#### 4. Does remission of sleep disorders predict remission of psychotic experiences?

This was investigated by logistic regression models testing if the remission of sleep disorders (i.e. present at baseline, absent at follow up) also predicted remission of psychotic experiences using logistic regression.

### Unregistered analyses

To additionally examine potential associations with distressing psychotic experiences, we completed the planned regression analyses with an alternative outcome: the presence/absence of at least one *distressing* psychotic experience. This followed further published analyses of the PQ-BC responses in the ABCD dataset that advised inclusion of distress to reduce false positives and thereby increase validity of the measure [30, 40].

In addition, we altered our analysis plan across all regression analyses based on ABCD analysis guidance included introduction of multi-level clustering by site and family [41], and use of population weights within the analyses.

## Results

### Demographic and descriptive results

Table 1 displays descriptive statistics for the study. Most descriptive variables remained relatively consistent between baseline and follow-up, however the follow-up group has a preponderance of White participants (59.5% compared to 52.1%) and fewer Black participants (9.3% at follow up compared to 15.0% at baseline).

**Table 1:** Demographic and descriptive variables.

| **Descriptive statistics** | **Baseline**<br>**n = 11830** | **12-month follow-up**<br>**n = 4910** |
| --- | --- | --- |
| Age – mean (SD) | 9y10m (7.4m) | 11y0m (7.6m) |
| Gender – n male (%) | 6162 (52.1%) | 2565 (52.2%) |
| Ethnicity – n(%) |  |  |
| White | 6161 (52.1%) | 2923 (59.5%) |
| Black | 1769 (15.0%) | 457 (9.3%) |
| Hispanic | 2391 (20.2%) | 930 (18.9%) |
| Asian | 250 (2.1%) | 115 (2.3%) |
| Other | 1238 (10.5%) | 485 (9.9%) |
| Socioeconomic status scale – mean (SD) | 0.47 (1.1) | 0.38 (1.0) |
| Child IQ Scaled Score – mean (SD) | 9.86 (3.0) | 10.14(2.9) |
| Neighbourhood deprivation percentile – mean (SD) | 39.22 (27.3) | 36.23 (25.3) |
| Family conflict scale – mean (SD) | 2.54 (2.0) | 2.47 (1.9) |
| Stimulant medication prescribed – n (%) | 722 (6.1%) | 322 (5.6%) |
| PQ-BC total – mean (SD) | 2.63 (3.6) | 1.71 (3.0) |
| PQ-BC distress – mean (SD) | 6.31 (10.6) | 4.05 (8.7) |
| SDSC total– mean (SD) | 36.53 (8.2) | 36.35 (7.9) |
| **Derived variable counts** |  |  |
| Sleep disorders present ($\geq 39$ cut off on SDSC) – n (%) | 3602 (30.4%) | 1392 (28.4%) |
| Sleep disorders onset (*absent at baseline, present at follow up*) – n(%) |  | 482 (9.8%) |
| Sleep disorders persist (*present at both baseline and follow up*) – n (%) |  | 909 (18.5%) |
| Sleep disorders remit (*present at baseline, absent at follow up*) – n (%) |  | 490 (10.0%) |
| Psychotic experiences present ($\geq 1$ on PQ-BC total) – n (%) | 7278 (61.5%) | 2298 (46.8%) |
| Psychotic experiences onset – n (%) |  | 473 (9.6%) |
| Psychotic experiences persist – n (%) |  | 1808 (36.8%) |
| Psychotic experiences remit – n (%) |  | 1208 (24.6%) |
| SDSC: Sleep Disorder Scale for Children, PQBC: Prodromal Questionnaire – Brief Child version ; SD = standard deviation |  |  |

Sleep disorders were present in the cohort at similar proportions at baseline and follow up (30.4% and 28.4% respectively). However, this masks some turnover, with roughly a third of the sleep disorder cases at follow-up representing ‘new onset’ cases (9.8%) to replace a similar proportion that remitted over the year (10.0%). For psychotic experiences there is a clear decline (61.5% of children reporting at baseline, versus 46.8% at follow-up), a result of the high remittance rate (24.6%) compared to new onset (9.6%). This is also reflected in the reduced average PQ-BC score at follow-up versus baseline.

### RQ1: Do sleep disorders and psychotic experiences co-occur in childhood?

The regression results associated with this research question can be found in Table 2. They illustrate that sleep disorders and psychotic experiences are significantly and strongly associated within the baseline time point. Sleep disorder symptoms above cut off is associated with an OR of 1.50 (95% CI 1.30-1.74) of at least one psychotic experience being present. The OR remains significant but reduces slightly (to 1.40, 95% CI 1.20-1.63) once the control variables are added (RQ1f). Results were similar in the additional analyses using presence of at least one distressing psychotic experience as the outcome in the unadjusted analysis (OR =1.49; 95% CI 1.29-1.71) and in the adjusted analysis (OR = 1.39, 95% CI 1.20-1.60). Tables with the full details of all analyses related to at least one distressing psychotic experiences are show in Supplementary Table 2.

**Table 2:** Regression analyses of co-occurrence of sleep disorders and psychotic experiences at baseline (T0)

| Model (outcome) | Parameters | B | 95% CI | p-value | AIC |
| --- | --- | --- | --- | --- | --- |
| Model 1a (PQBC total T0)<sup>a</sup> | SDSC total T0 | 0.048<sup>a</sup> | 0.04, 0.06 | <0.001 | 25850 |
| Model (outcome) | Parameters | Odds | 95% CI | p-value | AIC |
| Model 1b (PQBC cut-off T0)<sup>b</sup> | SDSC total T0 | 1.028 | 1.02, 1.04 | <0.001 | 6320 |
| Model 1c (PQBC cut-off T0)<sup>b</sup> | SDSC cut off T0 | 1.503 | 1.30, 1.74 | <0.001 | 6330 |
| Model (outcome) | Parameters | B | 95% CI | p-value | AIC |
| Model 1d (PQBC total T0)<sup>a</sup> | SDSC total T0 | 0.037 | 0.02, 0.05 | <0.001 | 24131 |
|  | Gender | -0.289 | -0.49, -0.09 | 0.004 |  |
|  | Ethnicity | 0.039 | -0.04, 0.12 | 0.347 |  |
|  | Socioeconomic status | 0.183 | 0.08, 0.29 | 0.001 |  |
|  | Neighbourhood deprivation | 0.011 | 0.01, 0.02 | <0.001 |  |
|  | IQ | -0.062 | -0.10, -0.03 | <0.001 |  |
|  | Family conflict | -0.003 | -0.06, 0.05 | 0.927 |  |
|  | Stimulant medication | 0.603 | 0.21, 1.00 | 0.003 |  |
| Model (outcome) | Parameters | Odds | 95% CI | p-value | AIC |
| Model 1e (PQBC cut-off T0)<sup>b</sup> | SDSC total T0 | 1.022 | 1.01, 1.03 | <0.001 | 5860 |
|  | Gender | 0.829 | 0.73, 0.94 | 0.005 |  |
|  | Ethnicity | 1.003 | 0.95, 1.06 | 0.897 |  |
|  | Socioeconomic status | 1.124 | 1.04, 1.21 | 0.003 |  |
|  | Neighbourhood deprivation | 1.009 | 1.01, 1.01 | <0.001 |  |
|  | IQ | 0.937 | 0.92, 0.96 | <0.001 |  |
|  | Family conflict | 0.998 | 0.96, 1.03 | 0.913 |  |
|  | Stimulant medication | 1.247 | 0.94, 1.65 | 0.121 |  |
| Model 1f (PQBC cut off T0)<sup>b</sup> | SDSC cut off T0 | 1.401 | 1.20, 1.63 | <0.001 | 5863 |
|  | Gender | 0.828 | 0.73, 0.94 | 0.004 |  |
|  | Ethnicity | 1.006 | 0.95, 1.06 | 0.826 |  |
|  | Socioeconomic status | 1.131 | 1.05, 1.22 | 0.001 |  |
|  | Neighbourhood deprivation | 1.009 | 1.01, 1.01 | <0.001 |  |
|  | IQ | 0.937 | 0.92, 0.96 | <0.001 |  |
|  | Family conflict | 1.003 | 0.97, 1.04 | 0.890 |  |
|  | Stimulant medication | 1.264 | 0.96, 1.67 | 0.098 |  |

<sup>a</sup> = linear regression; <sup>b</sup> = logistic regression; SDSC = Sleep Disorder Scale for Children; PQBC = Prodromal Questionnaire – Brief Child version; B = standardised beta; CI = confidence intervals; AIC = Akaike information criterion.

### RQ2: Do sleep disorders predict onset of psychotic experiences?

The results of the logistic regression analyses on this research question are shown in Table 3. Sleep disorder symptoms at baseline were found to predict psychotic experiences one year later (OR=1.62, 95% CI 1.41-1.86), even once sleep disorder symptoms at the later time point were controlled for (OR=1.41 95% CI 1.20-1.65). This association remained after potential confounding variables were added (OR=1.32, 95% CI 1.11-1.57). Again, results were similar when presence of distressing psychotic experiences was used as the outcome in both the unadjusted (OR = 1.39, 95% CI 1.18-1.64) and adjusted analysis (OR = 1.33, 95% CI 1.12-1.58)

**Table 3:** Regression analyses of whether sleep disorders at baseline predict onset of psychotic experiences at 12 months

| **Model (outcome)** | **Parameters** | **Odds** | **95% CI** | **p-value** | **AIC** |
| --- | --- | --- | --- | --- | --- |
| Model 2a (PQBC cut off T1) | SDSC cut off T0 | 1.621 | 1.41, 1.86 | <0.001 | 6548 |
| Model 2b (PQBC cut off T1) | SDSC cut off T0 | 1.407 | 1.20, 1.65 | <0.001 | 6538 |
|  | SDSC cut off T1 | 1.325 | 1.13, 1.56 | 0.001 |  |
| Model 2c (PQBC cut off T1) | SDSC cut off T0 | 1.496 | 1.29, 1.74 | <0.001 | 6048 |
|  | Gender | 0.799 | 0.70, 0.91 | 0.001 |  |
|  | Ethnicity | 1.031 | 0.98, 1.09 | 0.257 |  |
|  | Socioeconomic status | 1.171 | 1.09, 1.26 | <0.001 |  |
|  | Neighbourhood deprivation | 1.008 | 1.00, 1.01 | <0.001 |  |
|  | IQ | 0.959 | 0.94, 0.98 | <0.001 |  |
|  | Family conflict | 0.986 | 0.95, 1.02 | 0.430 |  |
|  | Stimulant medication | 1.649 | 1.26, 2.16 | <0.001 |  |
| Model 2d (PQBC cut off T1) | SDSC cut off T0 | 1.320 | 1.11, 1.57 | 0.001 | 6041 |
|  | SDSC cut off T1 | 1.294 | 1.09, 1.53 | 0.003 |  |
|  | Gender | 0.794 | 0.70, 0.91 | 0.001 |  |
|  | Ethnicity | 1.030 | 0.98, 1.09 | 0.267 |  |
|  | Socioeconomic status | 1.163 | 1.08, 1.25 | <0.001 |  |
|  | Neighbourhood deprivation | 1.008 | 1.00, 1.01 | <0.001 |  |
|  | IQ | 0.959 | 0.94, 0.98 | <0.001 |  |
|  | Family conflict | 0.983 | 0.95, 1.02 | 0.356 |  |
|  | Stimulant medication | 1.605 | 1.22, 2.10 | <0.001 |  |

SDSC = Sleep Disorder Scale for Children; PQBC = Prodromal Questionnaire – Brief Child version; CI = confidence intervals; AIC = Akaike information criterion

### RQ3: Do sleep disorders predict persistence of psychotic experiences?

The results of the logistic regression analyses relating to persistence of psychotic experiences are presented in Table 4. Presence of sleep disorder symptoms at baseline (OR=1.71, 95% CI 1.48-1.97) and persistence of sleep disorder symptoms (OR=1.89, 95% CI 1.60-2.23) were highly significant predictors of persistence of psychotic experiences at 12 months. These effects remained strong even once control variables were added (ORs of 1.56 and 1.72 for baseline and persisting sleep disorder symptoms respectively). The pattern of results was similar when presence of distressing psychotic experiences was used as the outcome variable (unadjusted OR=1.81, 95% CI 1.51-2.17, adjusted OR=1.62, 95% CI 1.34-1.97 for persistence of sleep disorder symptoms)

**Table 4:** Regression analyses of whether sleep disorders at baseline predict persistence of psychotic experiences at 12 months

| Model (outcome) | Parameters | Odds | 95% CI | p-value | AIC |
| --- | --- | --- | --- | --- | --- |
| Model 3a (PQBC persist) | SDSC cut off T0 | 1.707 | 1.48, 1.97 | <0.001 | 6192 |
| Model 3b (PQBC persist) | SDSC persist | 1.891 | 1.60, 2.23 | <0.001 | 6188 |
| Model 3c (PQBC persist) | SDSC cut off T0 | 1.562 | 1.34, 1.82 | <0.001 | 5710 |
|  | Gender | 0.798 | 0.70, 0.91 | 0.001 |  |
|  | Ethnicity | 0.999 | 0.95, 1.05 | 0.957 |  |
|  | Socioeconomic status | 1.163 | 1.08, 1.25 | <0.001 |  |
|  | Neighbourhood deprivation | 1.009 | 1.01, 1.01 | <0.001 |  |
|  | IQ | 0.953 | 0.93, 0.98 | <0.001 |  |
|  | Family conflict | 0.989 | 0.95, 1.03 | 0.553 |  |
|  | Stimulant medication | 1.471 | 1.13, 1.92 | 0.005 |  |
| Model 3d (PQBC persist) | SDSC persist | 1.716 | 1.44, 2.04 | <0.001 | 5706 |
|  | Gender | 0.796 | 0.70, 0.91 | 0.001 |  |
|  | Ethnicity | 0.998 | 0.95, 1.05 | 0.943 |  |
|  | Socioeconomic status | 1.160 | 1.08, 1.24 | <0.001 |  |
|  | Neighbourhood deprivation | 1.009 | 1.01, 1.01 | <0.001 |  |
|  | IQ | 0.952 | 0.93, 0.98 | <0.001 |  |
|  | Family conflict | 0.991 | 0.96, 1.03 | 0.627 |  |
|  | Stimulant medication | 1.460 | 1.12, 1.91 | 0.005 |  |

persist = above cut off at T0 and T1; SDSC = Sleep Disorder Scale for Children; PQBC = Prodromal Questionnaire – Brief Child version; CI = confidence intervals; AIC = Akaike information criterion.

### RQ4: Do sleep disorders predict remission of psychotic experiences?

Remission of sleep disorder symptoms was not a significant predictor of remission of psychotic experiences (OR=1.041, p=0.766, 95% CI 0.80, 1.35) in the uncontrolled model, therefore the analysis with control variables was not carried out. Again, results were similar when predicting distressing psychotic experiences as an outcome (OR=1.06, p=0.408, 95% CI 0.84-1.33, AIC=5123).

### Differences arising from weighted analysis

The results using the population weighted values are reported in Supplementary Material 3. The pattern of all results remained substantially the same except for non-white ethnicity which became a significant predictor of onset of psychotic experiences at 12 months.

## Discussion

The current study found that sleep disorder symptoms were strong and significant predictors of both the co-occurrence and persistence of psychotic experiences in 9-11 year olds, with associations remaining robust after controlling for potential confounders such as socioeconomic status, ethnicity, IQ, and stimulant medication. The prediction of persistence of psychotic experiences is especially relevant, given that the persistence of psychotic experiences through adolescence is associated with most severe mental health outcomes [42]. The remission of sleep disorders did not predict remission of psychotic experiences, and possible reasons for this negative association are discussed below. The pattern of results remained consistent when only distressing psychotic experiences were included, and when weighting was applied. Overall the results provide further support for a relationship between sleep problems and psychotic experiences during preadolescence.

Psychotic experiences were common in our study – reported by over two-thirds of participants at baseline. This is similar to previous studies in this age group [9], and the relatively high rate of remission versus new onset by the one year follow up is also in keeping with existing developmental research of psychotic experiences [27, 43]. Sleep disorder symptoms were also relatively common in the study group, with just under a third of the cohort having significant sleep issues, similar to the 30-40% reported in previous studies [44]. Guidelines on child sleep difficulties tend to advise that they ameliorate over time, contrary to our findings that around two-thirds do not (at least over a one-year observation period). There is evidence of increasing rates of child and adolescent sleep disorders over time [45] suggesting a need for additional support for parents and health services to address sleep difficulties.

The current study extends the known relationship between sleep and psychotic experiences to early adolescence and addressed potential confounders, yet mediating mechanisms require further investigation. A particularly relevant mechanism linking sleep and psychotic experiences is negative affect, which is typically found to either partially or totally mediate this relationship in adult studies [3, 6, 46, 47]. This and other potential mediators (such as affect dysregulation) should be investigated once further ABCD cohorts are available (or in other appropriate longitudinal designs), or ideally in manipulation studies to allow tests of the direction of influences over time in this age group.

The findings with respect to persistence of psychotic experiences are particularly relevant to clinical outcomes. As discussed above, the persistence of psychotic experiences across adolescence is associated with particularly poor outcomes [42, 48]. In addition to potential mediating mechanistic factors (not examined in this study) it seems highly plausible that sleep problems and psychotic experiences could interact and exacerbate each other – for example, distressing psychotic experiences may lead to disturbed sleep, which then lead to higher vulnerability for psychotic experiences the next day, as indicated by experience sampling studies in adults with psychotic disorders [49]. This potential causal model of sleep in contributing to psychotic experiences would support interventions on sleep to prevent persistence of psychotic experiences. These have been piloted in adolescents at-risk of psychosis, with initial indications that sleep treatment does improve psychotic experiences – but this is awaiting further examination in a follow-up trial currently underway [50, 51].

However, this current analysis did not support a relationship between the remission of sleep disorders and the remission of psychotic-experiences. As this study was observational, it is not possible to confirm a (lack of) causal relationship although several hypotheses can be generated. The first is that sleep disorders may cause increase and persistence in psychotic symptoms through altering a mediating mechanism that remains when sleep disorders resolve. For example, affective dysregulation has been highlighted as a potential mediating factor [52] with some evidence that this may also be apparent in children [18], as was also highlighted in Paper 1. Alternatively, sleep disorders may cause sensitisation of the mechanisms that generate psychotic symptoms but more rapidly than desensitisation occurs when sleep disorders resolve, and although recovery may occur, it may need more than the year interval measured with this data. Mechanisms here may be neurocognitive (for example, arousal-based increases in aberrant salience) or social (for example, sleep disorder causing behaviour difficulties, exclusion, or victimisation, subsequently impacting on psychotic experiences). Nevertheless, this does raise uncertainty about the potential efficacy of sleep improvement intervention in reducing psychotic symptoms in preadolescent children, given than an association between improved sleep and reduced psychotic experiences would be expected in this study if there were a simple causal association.

Besides our primary focus on the influence of sleep on psychotic experiences, our findings support the association of a wide range of epidemiological factors (male gender, socioeconomic status, neighbourhood deprivation) with psychotic experiences in this age group. Lower socioeconomic status of parents and increased neighbourhood deprivation were consistent predictors as supported by previous research [53]. Non-white ethnicity was not found to be a significant predictor in the unweighted models but was significant in some analyses once the models were weighted, supporting the longstanding association reported elsewhere [33]. Lastly, the strong relationship between stimulant medication and psychotic experiences is in keeping with previous literature [54].

### Limitations

We note that the observational nature of this study limits the causal conclusions we can draw as we cannot rule out that any changes or associations observed are due to unobserved factors. The current observation is also limited only to one year, which may be considered a short period of time to observe longitudinal associations between exposure and outcome – raising the possibility that the one-year associations are merely reflecting the concurrent links at baseline. In response to this we would point to the changes in sleep disorder and psychotic symptom occurrence over the study period as indicating change does occur in this time period, and therefore it is relevant to investigate the predictors of this variability.

We have been limited in variables available by the measures used in the ABCD study, which have their own benefits and limitations. For example, while self-reported scales for psychotic experiences in adolescence are recommended, false positives are known to occur [55, 56] and therefore these associations need to be additionally explored with alternative measures. The sleep measure was parent-reported and parents may vary in their detection of sleep disorders in their offspring, and some sleep disorders may be more likely to be observed by parents (e.g. sleep-walking versus insomnia).

In conclusion, this study indicates that sleep disorders in late childhood/early adolescence are common, and are associated with increased likelihood of reporting psychotic experiences both at the time and at one year follow up. Sleep disorders were also strongly associated with persistence of psychotic experiences from baseline to one year follow-up – this is of particular importance given that persistence of psychotic experiences in this period is associated with later psychotic disorder. These associations remained even while controlling for a broad range of confounding variables. This study suggests further attention towards sleep disorders in this period both as a plausible contributing factor to later mental health problems, but also as a possible preventative treatment target.

## Data Availability

The data are available from application the NIH ABCD study committee. Our pre-registered analysis plan and analysis script are available at https://osf.io/8ks72/

## Acknowledgements

Data used in the preparation of this article were obtained from the Adolescent Brain Cognitive Development^SM^ (ABCD) Study (https://abcdstudy.org), held in the NIMH Data Archive (NDA). This is a multisite, longitudinal study designed to recruit more than 10,000 9-10 year-olds and follow them over 10 years into early adulthood. The ABCD Study® is supported by the National Institutes of Health and additional federal partners under award numbers U01DA041022, U01DA041028, U01DA041048, U01DA041089, U01DA041106, U01DA041117, U01DA041120, U01DA041134, U01DA041148, U01DA041156, U01DA041174, U24DA041123, U24DA041147, U01DA041093, and U01DA041025. A full list of supporters is available at https://abcdstudy.org/federal-partners.html. A listing of participating sites and a complete listing of the study investigators can be found at https://abcdstudy.org/Consortium_Members.pdf. ABCD consortium investigators designed and implemented the study and/or provided data but did not necessarily participate in analysis or writing of this report. This manuscript reflects the views of the authors and may not reflect the opinions or views of the NIH or ABCD consortium investigators. The ABCD data repository grows and changes over time. The ABCD data used in this report came from doi: 10.15154/1521349. DOIs can be found at https://doi.org

## References

1. Reeve S, Sheaves B, Freeman D (2015) The role of sleep dysfunction in the occurrence of delusions and hallucinations: A systematic review. Clinical Psychology Review 42:96–115. https://doi.org/10.1016/j.cpr.2015.09.001

2. Waite F, Sheaves B, Isham L, et al (2019) Sleep and schizophrenia: From epiphenomenon to treatable causal target. Schizophrenia Research. https://doi.org/10.1016/J.SCHRES.2019.11.014

3. Reeve S, Emsley R, Sheaves B, Freeman D (2018) Disrupting Sleep: The Effects of Sleep Loss on Psychotic Experiences Tested in an Experimental Study With Mediation Analysis. Schizophrenia Bulletin 44:662–671. https://doi.org/10.1093/schbul/sbx103

4. Freeman D, Sheaves B, Goodwin GM, et al (2017) The effects of improving sleep on mental health (OASIS): a randomised controlled trial with mediation analysis. Lancet Psychiatry 4:749–758. https://doi.org/10.1016/S2215-0366(17)30328-0

5. Hennig T, Lincoln TM (2017) Sleeping Paranoia Away? An Actigraphy and Experience-Sampling Study with Adolescents. Child Psychiatry & Human Development 1–10. https://doi.org/10.1007/s10578-017-0729-9

6. Reeve S, Nickless A, Sheaves B, Freeman D (2018) Insomnia, negative affect, and psychotic experiences: Modelling pathways over time in a clinical observational study. Psychiatry Research 269:673–680. https://doi.org/10.1016/j.psychres.2018.08.090

7. Freeman D, Sheaves B, Waite F, et al (2020) Sleep disturbance and psychiatric disorders. The Lancet Psychiatry 7:628–637. https://doi.org/10.1016/S2215-0366(20)30136-X

8. Gradisar M, Gregory AM, Tikotzky L (2020) Is sleep the red flag to psychopathology’s bull? Journal of Child Psychology and Psychiatry and Allied Disciplines 61:1055–1057. https://doi.org/10.1111/jcpp.13331

9. Laurens KR, Hobbs MJ, Sunderland M, et al (2012) Psychotic-like experiences in a community sample of 8000 children aged 9 to 11 years: An item response theory analysis. Psychological Medicine 42:1495–1506. https://doi.org/10.1017/S0033291711002108

10. Linscott RJ, van Os J (2013) An updated and conservative systematic review and meta-analysis of epidemiological evidence on psychotic experiences in children and adults: on the pathway from proneness to persistence to dimensional expression across mental disorders. Psychological Medicine 43:1133–1149. https://doi.org/10.1017/S0033291712001626

11. McGrath JJ, Saha S, Al-Hamzawi A, et al (2015) Psychotic Experiences in the General Population. JAMA Psychiatry 72:697. https://doi.org/10.1001/jamapsychiatry.2015.0575

12. Maijer K, Hayward M, Fernyhough C, et al (2019) Hallucinations in Children and Adolescents: An Updated Review and Practical Recommendations for Clinicians. Schizophrenia Bulletin 45:S43–S55. https://doi.org/10.1093/schbul/sby119

13. Dominguez MDG, Wichers M, Lieb R, et al (2011) Evidence that onset of clinical psychosis is an outcome of progressively more persistent subclinical psychotic experiences: an 8-year cohort study. Schizophrenia Bulletin 37:84–93. https://doi.org/10.1093/schbul/sbp022

14. Fisher H, Caspi A, Poulton R (2013) Specificity of childhood psychotic symptoms for predicting schizophrenia by 38 years of age: a birth cohort study. Psychological Medicine 43:2077–86. https://doi.org/10.1017/S0033291712003091

15. Davies J, Sullivan S, Zammit S (2018) Adverse life outcomes associated with adolescent psychotic experiences and depressive symptoms. Soc Psychiatry Psychiatr Epidemiol 53:497–507. https://doi.org/10.1007/s00127-018-1496-z

16. Kelleher I, Keeley H, Corcoran P, et al (2012) Clinicopathological significance of psychotic experiences in non-psychotic young people: evidence from four population-based studies. The British Journal of Psychiatry 201:26–32. https://doi.org/10.1192/bjp.bp.111.101543

17. Trotta A, Arseneault L, Caspi A, et al (2020) Mental Health and Functional Outcomes in Young Adulthood of Children With Psychotic Symptoms: A Longitudinal Cohort Study. Schizophr Bull 46:261–271. https://doi.org/10.1093/schbul/sbz069

18. Jeppesen P, Clemmensen L, Munkholm A, et al (2014) Psychotic experiences co-occur with sleep problems, negative affect and mental disorders in preadolescence. Journal of child psychology and psychiatry, and allied disciplines 56:558–565. https://doi.org/10.1111/jcpp.12319

19. Koopman-Verhoeff ME, Bolhuis K, Cecil CAM, et al (2018) During day and night: Childhood psychotic experiences and objective and subjective sleep problems. Schizophrenia Research 206:127–134

20. Lee YJ, Cho S-J, Cho IH, et al (2012) The relationship between psychotic-like experiences and sleep disturbances in adolescents. Sleep medicine 13:1021–7. https://doi.org/10.1016/j.sleep.2012.06.002

21. Fisher H, Lereya ST, Thompson A, et al (2014) Childhood parasomnias and psychotic experiences at age 12 years in a United Kingdom birth cohort. Sleep 37:475–82. https://doi.org/10.5665/sleep.3478

22. Morales-Muñoz I, Broome MR, Marwaha S (2020) Association of Parent-Reported Sleep Problems in Early Childhood With Psychotic and Borderline Personality Disorder Symptoms in Adolescence. JAMA Psychiatry. https://doi.org/10.1001/jamapsychiatry.2020.1875

23. Thompson A, Lereya ST, Lewis G, et al (2015) Childhood sleep disturbance and risk of psychotic experiences at 18: UK birth cohort. British Journal of Psychiatry 207:23–29. https://doi.org/10.1192/bjp.bp.113.144089

24. Gregory AM, Sadeh A (2012) Sleep, emotional and behavioral difficulties in children and adolescents. Sleep Medicine Reviews 16:129–136. https://doi.org/10.1016/j.smrv.2011.03.007

25. Zhang J, Paksarian D, Lamers F, et al (2017) Sleep Patterns and Mental Health Correlates in US Adolescents. Journal of Pediatrics 182:137–143. https://doi.org/10.1016/j.jpeds.2016.11.007

26. Dong L, Gumport NB, Martinez AJ, Harvey AG (2019) Is improving sleep and circadian problems in adolescence a pathway to improved health? A mediation analysis. Journal of Consulting and Clinical Psychology 87:757–771. https://doi.org/10.1037/ccp0000423

27. Rubio JM, Sanjuán J, Flórez-Salamanca L, Cuesta MJ (2012) Examining the course of hallucinatory experiences in children and adolescents: A systematic review. Schizophrenia Research 138:248–254. https://doi.org/10.1016/j.schres.2012.03.012

28. Feldstein Ewing S, Luciana M (2018) The Adolescent Brain Cognitive Development (ABCD) Consortium: Rationale, Aims, and Assessment Strategy [Special Issue]. Developmental Cognitive Neuroscience 32:1–164

29. Volkow ND, Koob GF, Croyle RT, et al (2018) The conception of the ABCD study: From substance use to a broad NIH collaboration. Developmental Cognitive Neuroscience 32:4–7. https://doi.org/10.1016/j.dcn.2017.10.002

30. Karcher NR, Barch DM, Avenevoli S, et al (2018) Assessment of the prodromal questionnaire-brief child version for measurement of self-reported psychoticlike experiences in childhood. JAMA Psychiatry 75:853–861. https://doi.org/10.1001/jamapsychiatry.2018.1334

31. Bruni O, Ottaviano S, Guidetti V, et al (1996) The Sleep Disturbance Scale for Children (SDSC) construction and validation of an instrument to evaluate sleep disturbances in childhood and adolescence. Journal of Sleep Research 5:251–261. https://doi.org/10.1111/j.1365-2869.1996.00251.x

32. Biggs SN, Lushington K, Martin J, et al (2013) Gender, socioeconomic, and ethnic differences in sleep patterns in school-aged children. Sleep Medicine 14:1304–1309. https://doi.org/10.1016/j.sleep.2013.06.014

33. Jongsma HE, Gayer-Anderson C, Tarricone I, et al (2020) Social disadvantage, linguistic distance, ethnic minority status and first-episode psychosis: Results from the EU-GEI case-control study. Psychological Medicine. https://doi.org/10.1017/S003329172000029X

34. Kivimäki M, Batty GD, Pentti J, et al (2020) Association between socioeconomic status and the development of mental and physical health conditions in adulthood: a multi-cohort study. The Lancet Public Health 5:e140–e149. https://doi.org/10.1016/S2468-2667(19)30248-8

35. Tomfohr-Madsen L, Cameron EE, Dhillon A, et al (2020) Neighborhood socioeconomic status and child sleep duration: A systematic review and meta-analysis. Sleep Health: Journal of the National Sleep Foundation 6:550–562. https://doi.org/10.1016/j.sleh.2020.02.012

36. Gregory AM, Caspi A, Moffitt TE, Poulton R (2006) Family Conflict in Childhood: A Predictor of Later Insomnia. Sleep 29:1063–1067. https://doi.org/10.1093/sleep/29.8.1063

37. Cechnicki A, Bielańska A, Hanuszkiewicz I, Daren A (2013) The predictive validity of expressed emotions (EE) in schizophrenia. A 20-year prospective study. J Psychiatr Res 47:208–214. https://doi.org/10.1016/j.jpsychires.2012.10.004

38. Horwood J, Salvi G, Thomas K, et al (2008) IQ and non-clinical psychotic symptoms in 12-year-olds: results from the ALSPAC birth cohort. British Journal of Psychiatry 193:185–91. https://doi.org/10.1192/bjp.bp.108.051904

39. Kidwell KM, Dyk TRV, Lundahl A, Nelson TD (2015) Stimulant Medications and Sleep for Youth With ADHD: A Meta-analysis. Pediatrics 136:1144–1153. https://doi.org/10.1542/peds.2015-1708

40. Karcher NR, Loewy RL, Savill M, et al (2020) Replication of Associations With Psychotic-Like Experiences in Middle Childhood From the Adolescent Brain Cognitive Development (ABCD) Study. Schizophrenia Bulletin Open 1:. https://doi.org/10.1093/schizbullopen/sgaa009

41. Heeringa S, Berglund P (2020) A Guide for Population-based Analysis of the Adolescent Brain Cognitive Development (ABCD) Study Baseline Data. bioRxiv 2020.02.10.942011. https://doi.org/10.1101/2020.02.10.942011

42. Healy C, Campbell D, Coughlan H, et al (2018) Childhood psychotic experiences are associated with poorer global functioning throughout adolescence and into early adulthood. Acta Psychiatrica Scandinavica 138:26–34. https://doi.org/10.1111/acps.12907

43. de Leede-Smith S, Barkus E (2013) A comprehensive review of auditory verbal hallucinations: lifetime prevalence, correlates and mechanisms in healthy and clinical individuals. Frontiers in Human Neuroscience 7:. https://doi.org/10.3389/fnhum.2013.00367

44. Fricke-Oerkermann L, Plück J, Schredl M, et al (2007) Prevalence and Course of Sleep Problems in Childhood. Sleep 30:1371–1377. https://doi.org/10.1093/sleep/30.10.1371

45. Matricciani L, Olds T, Petkov J (2012) In search of lost sleep: Secular trends in the sleep time of school-aged children and adolescents. Sleep Medicine Reviews 16:203–211. https://doi.org/10.1016/j.smrv.2011.03.005

46. Freeman D, Stahl D, McManus S, et al (2012) Insomnia, worry, anxiety and depression as predictors of the occurrence and persistence of paranoid thinking. Social Psychiatry and Psychiatric Epidemiology 47:1195–203. https://doi.org/10.1007/s00127-011-0433-1

47. Sheaves B, Bebbington P, Guy M, et al (2016) Insomnia and hallucinations in the general population: Findings from the 2000 and 2007 British Psychiatric Morbidity Surveys. Psychiatry Research 241:141–146. https://doi.org/10.1016/j.psychres.2016.03.055

48. Healy C, Brannigan R, Dooley N, et al (2019) Childhood and adolescent psychotic experiences and risk of mental disorder: a systematic review and meta-analysis. Psychological Medicine 49:1589–1599. https://doi.org/10.1017/S0033291719000485

49. Mulligan LD, Haddock G, Emsley R, et al (2016) High resolution examination of the role of sleep disturbance in predicting functioning and psychotic symptoms in schizophrenia: A novel experience sampling study. Journal of Abnormal Psychology 125:788–797. https://doi.org/10.1037/abn0000180

50. Bradley J, Freeman D, Chadwick E, et al (2018) Treating Sleep Problems in Young People at Ultra-High Risk of Psychosis: A Feasibility Case Series. Behavioural and Cognitive Psychotherapy 46:276–291. https://doi.org/10.1017/S1352465817000601

51. Waite F, Kabir T, Johns L, et al (2020) Treating sleep problems in young people at ultra-high-risk of psychosis: Study protocol for a single-blind parallel group randomised controlled feasibility trial (SleepWell). BMJ Open 10:1–8. https://doi.org/10.1136/bmjopen-2020-045235

52. Akram U, Gardani M, Irvine K, et al (2020) Emotion dysregulation mediates the relationship between nightmares and psychotic experiences: results from a student population. npj Schizophrenia 6:1–7. https://doi.org/10.1038/s41537-020-0103-y

53. Solmi F, Lewis G, Zammit S, Kirkbride JB (2020) Neighborhood Characteristics at Birth and Positive and Negative Psychotic Symptoms in Adolescence: Findings from the ALSPAC Birth Cohort. Schizophrenia Bulletin 46:581–591. https://doi.org/10.1093/schbul/sbz049

54. Shyu Y-C, Yuan S-S, Lee S-Y, et al (2015) Attention-deficit/hyperactivity disorder, methylphenidate use and the risk of developing schizophrenia spectrum disorders: A nationwide population-based study in Taiwan. Schizophrenia Research 168:161–167. https://doi.org/10.1016/J.SCHRES.2015.08.033

55. Kelleher I, Harley M, Murtagh A, Cannon M (2011) Are Screening Instruments Valid for Psychotic-Like Experiences? A Validation Study of Screening Questions for Psychotic-Like Experiences Using In-Depth Clinical Interview. Schizophrenia Bulletin 37:362–369. https://doi.org/10.1093/schbul/sbp057

56. Lee K-W, Chan K-W, Chang W-C, et al (2016) A systematic review on definitions and assessments of psychotic-like experiences. Early Intervention in Psychiatry 10:3–16. https://doi.org/10.1111/eip.12228

