## Supplement 2 for "Sleep disorders predict the one-year onset, persistence, but not remission of psychotic experiences in pre-adolescence: a longitudinal analysis of the ABCD cohort data"

Supplementary Material 2: Distressing psychotic experiences as outcome variable

### Supplement 2 Table 1: Regression analyses of co-occurrence of sleep disorders and distressing psychotic experiences at baseline (T0)

| **Model (outcome)** | | | | | | | | | **Parameters** | **B** | **95% CI** | **p-value** | **AIC** |
| --- | --- | --- | --- | --- | --- | --- | --- | --- | --- | --- | --- | --- | --- |
| Model 1a (PQ-BC distress total T0) ^a^ | | | | | | | | | **SDSC total T0** | **0.143** | **0.11, 0.18** | **<0.001** | 36495 |
| **Model (outcome)** | | | | | | | | | **Parameters** | **Odds** | **95% CI** | **p-value** | **AIC** |
| Model 1b (PQ-BC cut-off T0) ^b^ | | | | | | | | | **SDSC total T0** | **1.027** | **1.02, 1.04** | **<0.001** | 6508 |
| Model 1c (PQ-BC cut-off T0) ^b^ | | | | | | | | | SDSC cut off T0 | 1.486 | 1.29, 1.71 | <0.001 | 6520 |
| **Model (outcome)** | | | | | | | | | **Parameters** | **B** | **95% CI** | **p-value** | **AIC** |
| Model 1d (PQ-BC total T0) ^a^ | | | | | | | | | **SDSC total T0** | **0.113** | **0.07, 0.15** | **<0.001** | 34168 |
| Gender_(RV=Male)_ | | | | | | | | | | -0.336 | -0.92, 0.25 | 0.262 |  |
| Ethnicity_(RV=White)_ | | | | | | | | | | 0.156 | -0.08, 0.40 | 0.202 |  |
| Socioeconomic status | | | | | | | | | | 0.579 | 0.27, 0.89  0.02, 0.05 | <0.001 |  |
| Neighbourhood deprivation | | | | | | | | | | 0.034 | 0.02, 0.05 | <0.001 |  |
|  | | | | | | | | | IQ | -0.219 | -0.32, -0.12 | <0.001 |  |
|  | | | | | | | | | Family conflict | 0.033 | -0.13, 0.20 | 0.692 |  |
| Stimulant medication_(RV=Not prescribed)_ | | | | | | | | | | 1.667 | 0.48, 2.85 | 0.006 |  |
| **Model (outcome)** | | | | | | | | | **Parameters** | **Odds** | **95% CI** | **p-value** | **AIC** |
| Model 1e (PQ-BC cut-off T0) ^b^ | | | | | | | | | **SDSC total T0** | **1.021** | **1.01, 1.03** | **<0.001** | 6048 |
| Gender_(RV=Male)_ | | | | | | | | | | 1.082 | 0.95, 1.23 | <0.001 |  |
| Ethnicity_(RV=White)_ | | | | | | | | | | 1.040 | 0.99, 1.09 | 0.231 |  |
| Socioeconomic status | | | | | | | | | | 1.108 | 1.03, 1.19 | 0.138 |  |
| Neighbourhood deprivation | | | | | | | | | | 1.006 | 1.00, 1.01 | 0.003 |  |
|  | | | | | | | | | IQ | 0.951  0.951  06 | 0.93, 0.97 | <0.001 |  |
|  | | | | | | | | | Family conflict | 1.014 | 0.98, 1.05 | 0.427 |  |
| Stimulant medication_(RV=Not prescribed)_ | | | | | | | | | | 1.378 | 1.06, 1.79 | 0.016 |  |
| Model 1f (PQ-BC cut-off T0) ^b^ | | | | | | | | | **SDSC total T0** | **1.385** | **1.20, 1.60** | **<0.001** | 6052 |
| Gender_(RV=Male)_ | | | | | | | | | | 1.081 | 0.95, 1.23 | 0.238 |  |
| Ethnicity_(RV=White)_ | | | | | | | | | | 1.043 | 0.99, 1.10 | 0.113 |  |
| Socioeconomic status | | | | | | | | | | 1.115 | 1.04, 1.19 | 0.002 |  |
| Neighbourhood deprivation | | | | | | | | | | 1.006 | 1.00, 1.01 | <0.001 |  |
|  | | | | | | | | | IQ | 0.950 | 0.93, 0.97 | <0.001 |  |
|  | | | | | | | | | Family conflict | 1.019 | 0.98, 1.05 | 0.294 |  |
| Stimulant medication_(RV=Not prescribed)_ | | | | | | | | | | 1.400 | 1.08, 1.82 | 0.011 |  |

^a^ = linear regression; ^b^ = logistic regression; ^c^reference category = Male

SDSC = Sleep Disorder Scale for Children; PQBC = Prodromal Questionnaire – Brief Child version; B = standardised beta; CI = confidence intervals; AIC = Akaike information criterion; RV = reference value.

### Supplement 2 Table 2: Regression analyses of whether sleep disorders at baseline predict onset of distressing psychotic experiences at 12 months

| **Model (outcome)** | **Parameters** | **Odds** | **95% CI** | **p-value** | **AIC** |
| --- | --- | --- | --- | --- | --- |
| Model 2a (PQ-BC cut off T1) | **SDSC cut off T0** | **1.643** | **1.42, 1.90** | **<0.001** | **5892** |
|  |  |  | 0.14 |  | 5880 |
| Model 2b (PQ-BC cut off T1) | **SDSC cut off T0** | **1.393** | **1.18, 1.64** | **<0.001** | **5880** |
|  | SDSC cut off T1 | 1.384 | 1.17, 1.63 | <0.001 |  |
|  |  |  |  |  | 5459 |
| Model 2c (PQ-BC cut off T1) | **SDSC cut off T0** | **1.523** | **1.31, 1.77** | **<0.001** | **5459** |
|  | Gender_(RV=Male)_ | 1.020 | 0.89, 1.17 | 0.772 |  |
|  | Ethnicity_(RV=White)_ | 1.038 | 0.98, 1.10 | 0.174 |  |
| Socioeconomic status | | 1.161 | 1.09, 1.24 | <0.001 |  |
| Neighbourhood deprivation | | 1.007 | 1.00, 1.01 | <0.001 |  |
|  | IQ | 0.964 | 0.94, 0.99 | 0.003 |  |
|  | Family conflict | 0.973 | 0.94, 1.01 | 0.140 |  |
| Stimulant medication_(RV=Not prescribed)_ | | 1.568 | 1.20, 2.04 | 0.001 |  |
| Model 2d (PQ-BC cut off T1) | **SDSC cut off T0** | **1.327** | **1.12, 1.58** | **0.001** | **5451** |
|  | SDSC cut off T1 | 1.326 | 1.12, 1.57 | 0.001 |  |
|  | Gender_(RV=Male)_ | 1.014 | 0.88, 1.16 | 0.843 |  |
|  | Ethnicity_(RV=White)_ | 1.037 | 0.98, 1.1 | 0.185 |  |
| Socioeconomic status | | 1.154 | 1.08, 1.24 | <0.001 |  |
| Neighbourhood deprivation | | 1.007 | 1.00, 1.01 | <0.001 |  |
|  | IQ | 0.964 | 0.94, 0.99 | 0.003 |  |
|  | Family conflict | 0.970 | 0.93, 1.01 | 0.108 |  |
| Stimulant medication_(RV=Not prescribed)_ | | 1.520 | 1.17, 1.98 | 0.002 |  |

SDSC = Sleep Disorder Scale for Children; PQ-BC = Prodromal Questionnaire – Brief Child version; CI = confidence intervals; AIC = Akaike information criterion; RV = reference value.

### Supplement 2 Table 3: Regression analyses of whether sleep disorders at baseline predict persistence of distressing psychotic experiences at 12 months

| **Model (outcome)** | **Parameters** | **Odds** | **95% CI** | **p-value** | **AIC** |
| --- | --- | --- | --- | --- | --- |
| Model 3a (PQ-BC persist) | **SDSC cut off T0** | **1.550** | **1.36, 1.76** | **<0.001** | **5146** |
| Model 3b (PQ-BC persist) | **SDSC persist** | **1.810** | **1.51, 2.17** | **<0.001** | **4869** |
| Model 3c (PQ-BC persist) | **SDSC cut off T0** | **1.743** | **1.47, 2.07** | **<0.001** | **4482** |
|  | Gender_(RV=Male)_ | 1.102 | 0.94, 1.29 | 0.221 |  |
|  | Ethnicity_(RV=White)_ | 1.028 | 0.97, 1.09 | 0.383 |  |
| Socioeconomic status | | 1.165 | 1.08, 1.25 | <0.001 |  |
| Neighbourhood deprivation | | 1.006 | 1.00, 1.01 | 0.002 |  |
|  | IQ | 0.945 | 0.92, 0.97 | <0.001 |  |
|  | Family conflict | 0.975 | 0.93, 1.02 | 0.239 |  |
| Stimulant medication_(RV=Not prescribed)_ | | 1.711 | 1.28, 2.28 | <0.001 |  |
| Model 3d (PQ-BC persist) | **SDSC persist** | **1.624** | **1.34, 1.97** | **<0.001** | **4499** |
|  | Gender_(RV=Male)_ | 1.100 | 0.94, 1.29 | 0.226 |  |
|  | Ethnicity_(RV=White)_ | 1.027 | 0.97, 1.09 | 0.403 |  |
| Socioeconomic status | | 1.170 | 1.09, 1.26 | <0.001 |  |
| Neighbourhood deprivation | | 1.006 | 1.00, 1.01 | 0.001 |  |
|  | IQ | 0.944 | 0.92, 0.97 | <0.001 |  |
|  | Family conflict | 0.982 | 0.94, 1.02 | 0.402 |  |
| Stimulant medication_(RV=Not prescribed)_ | | 1.749 | 1.31, 2.33 | <0.001 |  |

persist = above cut off at T0 and T1; SDSC = Sleep Disorder Scale for Children; PQ-BC = Prodromal Questionnaire – Brief Child version; CI = confidence intervals; AIC = Akaike information criterion; RV = reference value.
