## Supplement 3 for "Sleep disorders predict the one-year onset, persistence, but not remission of psychotic experiences in pre-adolescence: a longitudinal analysis of the ABCD cohort data"

**Supplement 3 – Weighted analysis results**

#### [Supplement 3 Table 1: Sample demographics and descriptive variables (weighted) 2](#_Toc80011466)

#### [Supplement 3 Table 2a: Regression analyses of co-occurrence of sleep disorders and psychotic experiences at baseline (T0) – weighting method A 3](#_Toc80011467)

#### [Supplement 3 Table 2b: Regression analyses of co-occurrence of sleep disorders and psychotic experiences at baseline (T0) – weighting method B 4](#_Toc80011468)

#### [Supplement 3 Table 3a: Regression analyses of whether sleep disorders at baseline predict onset of psychotic experiences at 12 months – weighting method A 5](#_Toc80011469)

#### [Supplement 3 Table 3b: Regression analyses of whether sleep disorders at baseline predict onset of psychotic experiences at 12 months – weighting method B 6](#_Toc80011470)

#### [Supplement 3 Table 4a: Regression analyses of whether sleep disorders at baseline predict persistence of psychotic experiences at 12 months – weighting method A 7](#_Toc80011471)

#### [Supplement 3 Table 4b: Regression analyses of whether sleep disorders at baseline predict persistence of psychotic experiences at 12 months – weighting method B 8](#_Toc80011472)

#### [Supplement 3 RQ4a: Regression analysis of whether remission of sleep problems predicts remission of psychotic experiences (weighted method A): 9](#_Toc80011473)

#### [Supplement 3 RQ4a: Regression analysis of whether remission of sleep problems predicts remission of psychotic experiences (weighted method B): 9](#_Toc80011474)

### Supplement 3 Table 1: Sample demographics and descriptive variables (weighted)

| **Descriptive statistics** | **Baseline**  **n = 11830** | **12-month follow-up**  **n = 4910** |
| --- | --- | --- |
| Age (months) – mean (SE) | 119.2 (0.3) | 132.2 (0.5) |
| Gender – male % (SE) | 51.13 (0.1) | 51.04 (0.9) |
| Ethnicity % (SE) |  |  |
| White | 52.34 (5.6) | 59.48 (6.3) |
| Black | 13.31 (2.5) | 7.61 (1.6) |
| Hispanic | 23.83 (6.2) | 22.50 (6.7) |
| Asian | 3.61 (0.9) | 4.02 (0.9) |
| Other | 6.89 (1.0) | 6.40 (1.3) |
| Socioeconomic status scale – mean (SE) | 0.56 (0.1) | 0.47 (0.1) |
| Child IQ – mean (SE) | 9.74 (0.1) | 10.03 (0.1) |
| Neighbourhood deprivation percentile – mean (SE) | 43.49 (3.8) | 40.5 (3.9) |
| Family conflict scale – mean (SE) | 2.52 (0.1) | 2.45 (0.06) |
| Stimulant medication prescribed – % (SE) | 6.41 (<0.0) | 7.00 (<0.0) |
| PQ-BC total – mean (SE) | 2.74 (0.3) | 1.82 (0.2) |
| SDSC total– mean (SE) | 36.78 (0.3) | 36.59 (0.3) |
| **Derived variable counts** | | |
| Sleep disorders present *(≥39 cut off on SDSC)* – % (SE) | 31.71 (1.5) | 29.15 (1.6) |
| Sleep disorders new onset *(absent at baseline, present at follow up)* – % (SE) | | 9.56 (0.1) |
| Sleep disorders persisting *(present at both baseline and follow up)* – % (SE) | | 19.50 (1.6) |
| Sleep disorders remitting *(present at baseline, absent at follow up)* – % (SE) | | 10.48 (0.8) |
| Psychotic experiences present *(≥1 on PQ-BC total)* – % (SE) | 63.08 (2.6) | 48.25 (2.8) |
| Psychotic experiences new onset – % (SE) | | 9.24 (0.9) |
| Psychotic experiences persisting – % (SE) | | 29.01 (3.0) |
| Psychotic experiences remitting – % (SE) | | 24.51 (1.1) |

SDSC = Sleep Disorder Scale for Children; PQBC = Prodromal Questionnaire – Brief Child version; SE = standard error;

### Supplement 3 Table 2a: Regression analyses of co-occurrence of sleep disorders and psychotic experiences at baseline (T0) – weighting method A

| **Model (outcome)** | | | | | | | | | **Parameters** | **B** | **95% CI** | **p-value** | **AIC** |
| --- | --- | --- | --- | --- | --- | --- | --- | --- | --- | --- | --- | --- | --- |
| Model 1a (PQBC total T0) ^a^ | | | | | | | | | SDSC total T0 | 0.051 | 0.04, 0.06 | <0.001 | 26047 |
| **Model (outcome)** | | | | | | | | | **Parameters** | **Odds** | **95% CI** | **p-value** | **AIC** |
| Model 1b (PQBC cut-off T0) ^b^ | | | | | | | | | SDSC total T0 | 1.034 | 1.02, 1.04 | <0.001 | 6053 |
|  | | | | | | | | |  |  |  | <0.001 |  |
| Model 1c (PQBC cut-off T0) ^b^ | | | | | | | | | SDSC cut off T0 | 1.614 | 1.37, 1.9 | <0.001 | 6067 |
| **Model (outcome)** | | | | | | | | | **Parameters** | **B** | **95% CI** | **p-value** | **AIC** |
| Model 1d (PQBC total T0) ^a^ | | | | | | | | | SDSC total T0 | 0.040 | 0.03, 0.05 | <0.001 | 24325 |
|  | | | | | | | | | Gender | -0.248 | -0.45, -0.05 | 0.014 |  |
|  | | | | | | | | | Ethnicity | 0.055 | -0.03, 0.14 | 0.220 |  |
| Socioeconomic status | | | | | | | | | | 0.171 | 0.07, 0.28 | 0.001 |  |
| Neighbourhood deprivation | | | | | | | | | | 0.011 | 0.01, 0.02 | <0.001 |  |
|  | | | | | | | | | IQ | -0.053 | -0.09, -0.02 | 0.003 |  |
|  | | | | | | | | | Family conflict | 0.011 | -0.04, 0.07 | 0.688 |  |
| Stimulant medication | | | | | | | | | | 0.581 | 0.18, 0.98 | 0.004 |  |
| **Model (outcome)** | | | | | | | | | **Parameters** | **Odds** | **95% CI** | **p-value** | **AIC** |
| Model 1e (PQBC cut-off T0) ^b^ | | | | | | | | | SDSC total T0 | 1.026 | 1.02, 1.04 | <0.001 | 5641 |
|  | | | | | | | | | Gender | 0.835 | 0.72, 0.97 | 0.021 |  |
|  | | | | | | | | | Ethnicity | 1.032 | 0.96, 1.11 | 0.402 |  |
| Socioeconomic status | | | | | | | | | | 1.155 | 1.06, 1.26 | 0.001 |  |
| Neighbourhood deprivation | | | | | | | | | | 1.010 | 1.01, 1.01 | <0.001 |  |
|  | | | | | | | | | IQ | 0.942 | 0.92, 0.97 | <0.001 |  |
|  | | | | | | | | | Family conflict | 1.004 | 0.96, 1.05 | 0.852 |  |
| Stimulant medication | | | | | | | | | | 1.208 | 0.88, 1.66 | 0.249 |  |
| Model 1f (PQBC cut off T0) ^b^ | | | | | | | | | SDSC cut off T0 | 1.470 | 1.23, 1.75 | <0.001 | 5647 |
|  | | | | | | | | | Gender | 0.834 | 0.72, 0.97 | 0.020 |  |
|  | | | | | | | | | Ethnicity | 1.034 | 0.96, 1.11 | 0.378 |  |
| Socioeconomic status | | | | | | | | | | 1.167 | 1.07, 1.27 | <0.001 |  |
| Neighbourhood deprivation | | | | | | | | | | 1.010 | 1.01, 1.01 | <0.001 |  |
|  | | | | | | | | | IQ | 0.942 | 0.92, 0.97 | <0.001 |  |
|  | | | | | | | | | Family conflict | 1.010 | 0.97, 1.05 | 0.638 |  |
| Stimulant medication | | | | | | | | | | 1.237 | 0.90, 1.70 | 0.192 |  |

^a^ = linear regression; ^b^ = logistic regression; SDSC = Sleep Disorder Scale for Children; PQBC = Prodromal Questionnaire – Brief Child version; B = standardised beta; CI = confidence intervals; AIC = Akaike information criterion.

Supplement 3 Table 2b: Regression analyses of co-occurrence of sleep disorders and psychotic experiences at baseline (T0) – weighting method B

| **Model (outcome)** | | | | | | | | | **Parameters** | **B** | **95% CI** | **p-value** | **AIC** |
| --- | --- | --- | --- | --- | --- | --- | --- | --- | --- | --- | --- | --- | --- |
| Model 1a (PQBC total T0) ^a^ | | | | | | | | | SDSC total T0 | 0.051 | 0.04, 0.06 | <0.001 | 26054 |
| **Model (outcome)** | | | | | | | | | **Parameters** | **Odds** | **95% CI** | **p-value** | **AIC** |
| Model 1b (PQBC cut-off T0) ^b^ | | | | | | | | | SDSC total T0 | 1.029 | 1.02, 1.04 | <0.001 | 5027 |
|  | | | | | | | | |  |  |  | <0.001 |  |
| Model 1c (PQBC cut-off T0) ^b^ | | | | | | | | | SDSC cut off T0 | 1.501 | 1.29, 1.75 | <0.001 | 5039 |
| **Model (outcome)** | | | | | | | | | **Parameters** | **B** | **95% CI** | **p-value** | **AIC** |
| Model 1d (PQBC total T0) ^a^ | | | | | | | | | SDSC total T0 | 0.040 | 0.03, 0.05 | <0.001 | 24333 |
|  | | | | | | | | | Gender | -0.247 | -0.45, -0.05 | 0.015 |  |
|  | | | | | | | | | Ethnicity | 0.057 | -0.03, 0.15 | 0.207 |  |
| Socioeconomic status | | | | | | | | | | 0.170 | 0.07, 0.27 | 0.001 |  |
| Neighbourhood deprivation | | | | | | | | | | 0.011 | 0.01, 0.02 | <0.001 |  |
|  | | | | | | | | | IQ | -0.053 | -0.09, -0.02 | 0.003 |  |
|  | | | | | | | | | Family conflict | 0.011 | -0.04, 0.07 | 0.694 |  |
| Stimulant medication | | | | | | | | | | 0.582 | 0.18, 0.98 | 0.004 |  |
| **Model (outcome)** | | | | | | | | | **Parameters** | **Odds** | **95% CI** | **p-value** | **AIC** |
| Model 1e (PQBC cut-off T0) ^b^ | | | | | | | | | SDSC total T0 | 1.022 | 1.01, 1.03 | <0.001 | 4687 |
|  | | | | | | | | | Gender | 0.876 | 0.76, 1.01 | 0.066 |  |
|  | | | | | | | | | Ethnicity | 1.037 | 0.97, 1.11 | 0.296 |  |
| Socioeconomic status | | | | | | | | | | 1.147 | 1.06, 1.24 | 0.001 |  |
| Neighbourhood deprivation | | | | | | | | | | 1.008 | 1.00, 1.01 | <0.001 |  |
|  | | | | | | | | | IQ | 0.952 | 0.93, 0.98 | <0.001 |  |
|  | | | | | | | | | Family conflict | 1.002 | 0.96, 1.04 | 0.910 |  |
| Stimulant medication | | | | | | | | | | 1.151 | 0.86, 1.55 | 0.353 |  |
| Model 1f (PQBC cut off T0) ^b^ | | | | | | | | | SDSC cut off T0 | 1.374 | 1.17, 1.62 | <0.001 | 4693 |
|  | | | | | | | | | Gender | 0.875 | 0.76, 1.01 | 0.063 |  |
|  | | | | | | | | | Ethnicity | 1.038 | 0.97, 1.11 | 0.280 |  |
| Socioeconomic status | | | | | | | | | | 1.157 | 1.07, 1.25 | <0.001 |  |
| Neighbourhood deprivation | | | | | | | | | | 1.008 | 1.00, 1.01 | <0.001 |  |
|  | | | | | | | | | IQ | 0.952 | 0.93, 0.98 | <0.001 |  |
|  | | | | | | | | | Family conflict | 1.008 | 0.97, 1.05 | 0.693 |  |
| Stimulant medication | | | | | | | | | | 1.180 | 0.88, 1.59 | 0.274 |  |

^a^ = linear regression; ^b^ = logistic regression; SDSC = Sleep Disorder Scale for Children; PQBC = Prodromal Questionnaire – Brief Child version; B = standardised beta; CI = confidence intervals; AIC = Akaike information criterion.

### Supplement 3 Table 3a: Regression analyses of whether sleep disorders at baseline predict onset of psychotic experiences at 12 months – weighting method A

| **Model (outcome)** | **Parameters** | **Odds** | **95% CI** | **p-value** | **AIC** |
| --- | --- | --- | --- | --- | --- |
| Model 2a (PQBC cut off T1) | SDSC cut off T0 | 1.719 | 1.47, 2.01 | <0.001 | 6303 |
|  |  |  | 0.14 | <0.001 |  |
| Model 2b (PQBC cut off T1) | SDSC cut off T0 | 1.482 | 1.24, 1.77 | <0.001 | 6294 |
|  | SDSC cut off T1 | 1.346 | 1.12, 1.61 | 0.001 |  |
| Model 2c (PQBC cut off T1) | SDSC cut off T0 | 1.511 | 1.29, 1.77 | <0.001 | 5826 |
|  | Gender | 0.808 | 0.7, 0.93 | 0.003 |  |
|  | Ethnicity | 1.066 | 1.00, 1.13 | 0.041 |  |
| Socioeconomic status | | 1.231 | 1.14, 1.33 | <0.001 |  |
| Neighbourhood deprivation | | 1.009 | 1.01, 1.01 | <0.001 |  |
|  | IQ | 0.969 | 0.95, 0.99 | 0.013 |  |
|  | Family conflict | 0.994 | 0.96, 1.03 | 0.749 |  |
| Stimulant medication | | 1.702 | 1.28, 2.26 | <0.001 |  |
| Model 2d (PQBC cut off T1) | SDSC cut off T0 | 1.318 | 1.32, 1.32 | <0.001 | 5819 |
|  | SDSC cut off T1 | 1.328 | 1.33, 1.33 | <0.001 |  |
|  | Gender | 0.798 | 0.80, 0.80 | <0.001 |  |
|  | Ethnicity | 1.064 | 1.06, 1.07 | <0.001 |  |
| Socioeconomic status | | 1.220 | 1.22, 1.22 | <0.001 |  |
| Neighbourhood deprivation | | 1.009 | 1.01, 1.01 | <0.001 |  |
|  | IQ | 0.969 | 0.97, 0.97 | <0.001 |  |
|  | Family conflict | 0.992 | 0.99, 0.99 | <0.001 |  |
| Stimulant medication | | 1.677 | 1.67, 1.68 | <0.001 |  |

SDSC = Sleep Disorder Scale for Children; PQBC = Prodromal Questionnaire – Brief Child version; CI = confidence intervals; AIC = Akaike information criterion

Supplement 3 Table 3b: Regression analyses of whether sleep disorders at baseline predict onset of psychotic experiences at 12 months – weighting method B

| **Model (outcome)** | **Parameters** | **Odds** | | **95% CI** | **p-value** | **AIC** |
| --- | --- | --- | --- | --- | --- | --- |
| Model 2a (PQBC cut off T1) | SDSC cut off T0 | 1.568 | | 1.36, 1.81 | <0.001 | 5226 |
|  |  |  | | 0.14 |  |  |
| Model 2b (PQBC cut off T1) | SDSC cut off T0 | 1.391 | | 1.18, 1.64 | <0.001 | 5220 |
|  | SDSC cut off T1 | 1.268 | | 1.07, 1.50 | 0.006 |  |
| Model 2c (PQBC cut off T1) | SDSC cut off T0 | 1.384 | | 1.18, 1.62 | <0.001 | 4836 |
|  | Gender | 0.862 | | 0.75, 0.99 | 0.035 |  |
|  | Ethnicity | 1.072 | | 1.00, 1.15 | 0.039 |  |
| Socioeconomic status | | 1.214 | | 1.13, 1.30 | <0.001 |  |
| Neighbourhood deprivation | | 1.007 | | 1.00, 1.01 | <0.001 |  |
|  | IQ | 0.975 | | 0.95, 1.00 | 0.049 |  |
|  | Family conflict | 0.998 | | 0.96, 1.04 | 0.906 |  |
| Stimulant medication | | 1.583 | | 1.19, 2.10 | 0.001 |  |
| Model 2d (PQBC cut off T1) | SDSC cut off T0 | 1.248 | | 1.04, 1.49 | 0.015 | 4832 |
|  | SDSC cut off T1 | 1.242 | | 1.04, 1.48 | 0.017 |  |
|  | Gender | 0.857 | | 0.75, 0.98 | 0.029 |  |
|  | Ethnicity | 1.071 | | 1.00, 1.14 | 0.042 |  |
| Socioeconomic status | | 1.205 | | 1.12, 1.30 | <0.001 |  |
| Neighbourhood deprivation | | 1.007 | | 1.00, 1.01 | <0.001 |  |
|  | IQ | 0.975 | | 0.95, 1.00 | 0.045 |  |
|  | Family conflict | 0.995 | | 0.96, 1.03 | 0.798 |  |
| Stimulant medication | | 1.546 | 1.16, 2.05 | | 0.003 |  |

SDSC = Sleep Disorder Scale for Children; PQBC = Prodromal Questionnaire – Brief Child version; CI = confidence intervals; AIC = Akaike information criterion

### Supplement 3 Table 4a: Regression analyses of whether sleep disorders at baseline predict persistence of psychotic experiences at 12 months – weighting method A

| **Model (outcome)** | **Parameters** | **Odds** | **95% CI** | **p-value** | **AIC** |
| --- | --- | --- | --- | --- | --- |
| Model 3a (PQBC persist) | SDSC cut off T0 | 1.841 | 1.57, 2.16 | <0.001 | 5996 |
| Model 3b (PQBC persist) | SDSC persist | 2.058 | 1.71, 2.48 | <0.001 | 5994 |
| Model 3c (PQBC persist) | SDSC cut off T0 | 1.598 | 1.60, 1.60 | <0.001 | 5537 |
|  | Gender | 0.808 | 0.81, 0.81 | <0.001 |  |
|  | Ethnicity | 1.022 | 1.02, 1.02 | <0.001 |  |
| Socioeconomic status | | 1.220 | 1.22, 1.22 | <0.001 |  |
| Neighbourhood deprivation | | 1.009 | 1.01, 1.01 | <0.001 |  |
|  | IQ | 0.960 | 0.96, 0.96 | <0.001 |  |
|  | Family conflict | 1.002 | 1.00, 1.00 | <0.001 |  |
| Stimulant medication | | 1.508 | 1.51, 1.51 | <0.001 |  |
| Model 3d (PQBC persist) | SDSC persist | 1.795 | 1.46, 2.20 | <0.001 | 5532 |
|  | Gender | 0.813 | 0.69, 0.95 | 0.010 |  |
|  | Ethnicity | 1.025 | 0.95, 1.10 | 0.517 |  |
| Socioeconomic status | | 1.217 | 1.12, 1.32 | <0.001 |  |
| Neighbourhood deprivation | | 1.010 | 1.01, 1.01 | <0.001 |  |
|  | IQ | 0.963 | 0.94, 0.99 | 0.008 |  |
|  | Family conflict | 1.004 | 0.96, 1.05 | 0.844 |  |
| Stimulant medication | | 1.567 | 1.15, 2.14 | 0.005 |  |

persist = above cut off at T0 and T1; SDSC = Sleep Disorder Scale for Children; PQBC = Prodromal Questionnaire – Brief Child version; CI = confidence intervals; AIC = Akaike information criterion.

Supplement 3 Table 4b: Regression analyses of whether sleep disorders at baseline predict persistence of psychotic experiences at 12 months – weighting method B

| Model (outcome) | Parameters | Odds | 95% CI | p-value | AIC |
| --- | --- | --- | --- | --- | --- |
| Model 3a (PQBC persist) | SDSC cut off T0 | 1.671 | 1.44, 1.94 | <0.001 | 4989 |
| Model 3b (PQBC persist) | SDSC persist | 1.828 | 1.54, 2.17 | <0.001 | 4987 |
| Model 3c (PQBC persist) | SDSC cut off T0 | 1.473 | 1.25, 1.73 | <0.001 | 4609 |
|  | Gender | 0.862 | 0.75, 1.00 | 0.043 |  |
|  | Ethnicity | 1.042 | 0.97, 1.12 | 0.237 |  |
| Socioeconomic status | | 1.208 | 1.13, 1.30 | <0.001 |  |
| Neighbourhood deprivation | | 1.007 | 1.00, 1.01 | <0.001 |  |
|  | IQ | 0.969 | 0.94, 0.99 | 0.017 |  |
|  | Family conflict | 1.006 | 0.97, 1.05 | 0.744 |  |
| Stimulant medication | | 1.407 | 1.06, 1.86 | 0.017 |  |
| Model 3d (PQBC persist) | SDSC persist | 1.598 | 1.33, 1.92 | <0.001 | 4321 |
|  | Gender | 0.862 | 0.75, 0.99 | 0.042 |  |
|  | Ethnicity | 1.043 | 0.97, 1.12 | 0.232 |  |
| Socioeconomic status | | 1.203 | 1.12, 1.29 | <0.001 |  |
| Neighbourhood deprivation | | 1.007 | 1.00, 1.01 | <0.001 |  |
|  | IQ | 0.969 | 0.94, 0.99 | 0.016 |  |
|  | Family conflict | 1.007 | 0.97, 1.05 | 0.717 |  |
| Stimulant medication | | 1.406 | 1.06, 1.86 | 0.018 |  |

persist = above cut off at T0 and T1; SDSC = Sleep Disorder Scale for Children; PQBC = Prodromal Questionnaire – Brief Child version; CI = confidence intervals; AIC = Akaike information criterion.

### Supplement 3 RQ4a: Regression analysis of whether remission of sleep problems predicts remission of psychotic experiences (weighted method A):

Remission of sleep disorder symptoms was not a significant predictor of remission of psychotic experiences (OR=1.041, p=0.588, 95% CI 0.80, 1.35, p=0.766, AIC=5241) in the uncontrolled model, therefore the analysis with control variables was not carried out.

### Supplement 3 RQ4a: Regression analysis of whether remission of sleep problems predicts remission of psychotic experiences (weighted method B):

Remission of sleep disorder symptoms was not a significant predictor of remission of psychotic experiences (OR=1.011, p=0.588, 95% CI 0.79, 1.29, p-0.929) in the uncontrolled model, therefore the analysis with control variables was not carried out.
