## Supplement 1 for "Sleep disorders predict the one-year onset, persistence, but not remission of psychotic experiences in pre-adolescence: a longitudinal analysis of the ABCD cohort data"

| (Morgan et al., undefined/ed)(Reeve et al., 2015; Waite et al., 2019)**Package** | **Version** | **Citation** |
| --- | --- | --- |
| dplyr | 1.0.2 | (Wickham et al., 2020) |
| readr | 1.3.1 | (Wickham et al., 2018) |
| tidyr | 1.1.1 | (Wickham, 2020) |
| psych | 1.9.12.31 | (Revelle, 2020) |
| knitr | 1.29 | (Xie, 2020) |
| survey | 3.37 | (Lumley, 2020) |
| lme4 | 1.1.21 | (Bates et al., 2019) |
| lmertest | 3.1.1 | (Kuznetsova et al., 2019) |
| memisc | 0.99.22 | (Elff, 2020) |
| mfx | 1.2.2 | (Fernihough, 2019) |
| MASS | 7.3.51.5 | (Ripley, 2019) |
| parameters | 0.8.2 | (Lüdecke et al., 2019) |

**Supplementary document 1: R packages used in analysis**

Bates, D., Maechler, M., Bolke, B., & Walker, S. (2019). Linear Mixed-Effects Models using “Eigen” and S4 [R package lme4 version 1.1.21]. Comprehensive R Archive Network (CRAN).

Elff, M. (2020). Management of Survey Data and Presentation of Analysis Results [R package memisc version 0.99.22]. Comprehensive R Archive Network (CRAN).

Fernihough, A. (2019). Marginal Effects, Odds Ratios and Incidence Rate Ratios for GLMs [R package mfx version 1.2-2]. Comprehensive R Archive Network (CRAN).

Kuznetsova, A., Brockhoff, P., & Christensen, R. (2019). Tests in Linear Mixed Effects Models [R package lmerTest version 3.1.1]. Comprehensive R Archive Network (CRAN).

Lüdecke, D., Waggoner, P., & Makowski, D. (2019). A Unified Interface to Access Information from Model Objects in R [R package parameters version 0.8.2]. Journal of Open Source Software, 4(38), 1412. https://doi.org/10.21105/joss.01412

Lumley, T. (2020). survey: analysis of complex survey samples [R package survey version 3.37]. Comprehensive R Archive Network (CRAN).

Revelle, W. (2020). Procedures for Psychological, Psychometric, and Personality Research [R package psych version 1.9.12.31]. Comprehensive R Archive Network (CRAN).

Ripley, B. (2019). Support Functions and Datasets for Venables and Ripley’s MASS [R package MASS version 7.3.51.5]. Comprehensive R Archive Network (CRAN).

Wickham, H. (2020). Tidy Messy Data [R package tidyr version 1.1.1]. Comprehensive R Archive Network (CRAN).

Wickham, H., François, R., Henry, L., & Müller, K. (2020). A Grammar of Data Manipulation [R package dplyr version 1.0.2]. Comprehensive R Archive Network (CRAN).

Wickham, H., Hester, J., & Francois, R. (2018). Read Rectangular Text Data [R package readr version 1.3.1]. Comprehensive R Archive Network (CRAN).

Xie, Y. (2020). A General-Purpose Package for Dynamic Report Generation in R [R package knitr version 1.29]. Comprehensive R Archive Network (CRAN).
